# The contribution of white matter pathology, hypoperfusion, lesion load, and stroke recurrence to language deficits following acute subcortical left hemisphere stroke

**DOI:** 10.1101/2022.03.22.22272738

**Authors:** Massoud S. Sharif, Emily B. Goldberg, Alexandra Walker, Argye E. Hillis, Erin L. Meier

**Author notes:** **Corresponding author:** Erin L. Meier, Ph.D., CCC-SLP, Department of Neurology, Johns Hopkins University, School of Medicine, 600 N. Wolfe Street, Phipps 546C, Baltimore, MD 21287.

## Abstract

Aphasia, the loss of language ability following damage to the brain, is among the most disabling and common consequences of stroke. Subcortical stroke, occurring in the basal ganglia, thalamus, and/or deep white matter can result in aphasia, often characterized by word fluency, motor speech output, or sentence generation impairments. The link between greater lesion volume and acute aphasia is well documented, but the independent contributions of lesion location, cortical hypoperfusion, prior stroke, and white matter degeneration (leukoaraiosis) remain unclear, particularly in subcortical aphasia. Thus, we aimed to disentangle the contributions of each factor on language impairments in left hemisphere acute subcortical stroke survivors. Eighty patients with acute left hemisphere subcortical stroke (less than 10 days post-onset) participated. We manually traced acute lesions on diffusion-weighted scans and prior lesions on T2-weighted scans. Leukoaraiosis was rated on T2-weighted scans using the Fazekas et al. (1987) scale. Fluid-attenuated inversion recovery (FLAIR) scans were evaluated for hyperintense vessels in each vascular territory, providing an indirect measure of hypoperfusion in lieu of perfusion-weighted imaging. Compared to subcortical stroke patients without aphasia, patients with aphasia had greater acute and total lesion volume, were older, and had significantly greater damage to the internal capsule (which did not survive controlling for total lesion volume). Patients with aphasia did not differ from non-aphasic patients by other demographic or stroke variables. Age was the only significant predictor of aphasia status in a logistic regression model. Further examination of three participants with severe language impairments suggests that their deficits result from impairment in domain-general, rather than linguistic, processes. Given the variability in language deficits and imaging markers associated with such deficits, it seems likely that subcortical aphasia is a heterogeneous clinical syndrome with distinct causes across individuals.

## 1. Introduction

Aphasia, the loss of language ability following injury to the brain, is a common and often disabling consequence of acute left hemisphere (LH) cortical stroke (Berthier, 2005; Flowers et al., 2016; Tsouli et al., 2009). Aphasia has been reported in frequencies as high as 62% in acute ischemic stroke — a higher rate than in hemorrhagic stroke or in ischemic stroke at any other time point (Flowers et al., 2016).

The neural substrates of cortical aphasia are well-established (Plowman et al., 2012; Yassi et al., 2015). Subcortical aphasia, which occurs after strokes in the basal ganglia, thalamus, or deep white matter, is not as well characterized (Bouvier et al., 2017; Kang et al., 2017; Radanovic & Mansur, 2017). Subcortical aphasia lacks some of the clinical features of classic aphasia subtypes (Damasio et al., 1982), and the type and severity of language impairments after subcortical stroke varies widely (Bouvier et al., 2017; Caplan et al., 1990) — with no characteristic error pattern found among these patients generally (Kennedy & Murdoch, 1993). Whether subcortical structures have a direct role in processing language is controversial (Murdoch, 2001). The basal ganglia may be involved in the formulation and selection of language segments or merely in the motoric aspects of their release (Nadeau & Crosson, 1997). Similarly, the thalamus may be crucial to integrative language functions or may only support the attentional mechanisms that underlie them (Nadeau & Crosson, 1997). Here, further historical context is appropriate.

Nearly a century and a half ago, Broadbent believed the cells of the striatum store pre-formed articulatory patterns for spoken words (Broadbent, 1872). Others maintained that the basal ganglia mediated purely motoric functions (Kussmaul, 1877), and the idea that subcortical aphasia was merely the result of disconnecting cortical language areas emerged as well (Wernicke, 1874). In the mid-twentieth century, before the emergence of modern imaging, speech disturbances during intraoperative stimulation or ablation of the thalamus and basal ganglia in Parkinsonian patients continued this line of inquiry (Allan et al., 1966; Svennilson et al., 1960). More recently, a study of language in Huntington’s disease patients suggested that the contributions of the striatum to syntax processing are not merely a reflection of its role in working memory (Sambin et al., 2012). In a recent review, however, Radanovic & Mansur (2017) argue that neurodegenerative diseases such as Parkinson’s and Huntington’s, while both diseases of the basal ganglia, are imperfect models for subcortical lesions. They explain that language impairments are better attributed to the diffuse effect of these pathologies on the cerebral cortex.

Evidence from subcortical stroke itself has also contributed to this debate. Kuljic-Obradovic (2003), in a study of 32 aphasic patients with LH subcortical strokes, proposed a division of subcortical aphasia into three syndromes. In the first two, striatocapsular aphasia and aphasia associated with periventricular white matter lesions, he proposed patients display similar patterns of impairment in speech fluency and shortened phrase length with preserved comprehension and naming. Individuals with thalamic aphasia, on the other hand, present with fluent output but impaired comprehension and naming. Kuljic-Obradovic (2003) found that repetition is generally spared across these proposed subtypes. This dissociation, the author argues, suggests a phonetic processing role for the basal ganglia and a lexical-semantic processing role for the thalamus. In addition, Crosson et al. (2005) found that the left basal ganglia may have a role in reorganizing language production faculties to the right hemisphere after ischemic LH stroke.

More recent work has provided evidence that subcortical structures only indirectly support language. Bohsali & Crosson (2016) discuss two major functional loops through the basal ganglia and thalamus that support lexical selection and articulation. The first of these loops connects the left pre-supplementary motor area (SMA), dorsal caudate, and ventral anterior thalamus, while the second connects Broca’s area to the basal ganglia. The authors implicate the first loop in the retrieval of words from pre-existing lexical stores. Here, the role of the basal ganglia is to refine contrasts between desired and non-desired words in order to increase the signal-to-noise ratio (Bohsali & Crosson, 2016). Similarly, the second loop refines the contrast between desired and competing phonological/articulatory representations. Together, the two loops ensure the accurate selection and proper articulation of the desired lexical phrases. This explanation accords with a recent review by Nadeau (2021), who emphasized the strictly computational and non-data-specific functions of the basal ganglia in assisting language. The basal ganglia, Nadeau continues, do not have strict language functionalities, with the possible exception of representing movement verbs. Further, a fascinating review by Shi & Zhang (2020) on music therapy as treatment for aphasia suggests the basal ganglia facilitate language only insofar as they handle rhythm and beat processing, temporal prediction, and the execution of motor programs. As with the basal ganglia, Nadeau (2021) emphasized the purely computational nature of thalamic circuits involving language centers. The most likely explanation of thalamic aphasia, he writes, is diaschisis. Crosson (2019) posits that cortico-thalamo-cortical connections maintain semantic representations while they are compared to lexical items generated in language cortex. This comparison by the thalamus generates an error signal that allows the interface between semantic and lexical mechanisms to be fine-tuned until the desired match is achieved and eventually sent to other cortical areas.

The variation in the type and severity of language deficits after subcortical stroke is likely due to a variety of factors. The link between greater lesion volume and poorer language abilities in acute *cortical* aphasia has been consistently reported (Plowman et al., 2012), but such a link has only seen some support in subcortical aphasia patient populations (Mega & Alexander, 1994). In acute left hemisphere stroke survivors with a history of prior stroke, Goldberg et al. (2021) found that the total amount of brain damage incurred by strokes was a stronger predictor of acute language impairments than a categorical stroke history variable (i.e., history versus no history of prior stroke). Olsen et al. (1986), however, argued that infarcted subcortical tissue does not itself account for subcortical aphasia. They instead implicated cortical hypoperfusion (i.e., the deprivation of oxygenated blood flow to cerebral cortex due to artery occlusion) (Hillis et al., 2006) as the primary driver of language impairment after subcortical stroke. Further, Hillis et al. (2002) demonstrated that the reversal of cortical hypoperfusion is associated with the resolution of aphasia. They also reported that cortical hypoperfusion better predicted presence of aphasia than non-thalamic subcortical lesion volume did. The case for hypoperfusion as the primary cause of subcortical aphasia was strengthened by another study by Hillis et al. (2004), who found that aphasia classifications (Broca’s, Wernicke’s, Global, etc.) were linked to hypoperfusion in specific regions of the cerebral cortex. More recently, however, Sebastian et al. (2014) found that aphasia can occur without hypoperfusion in left thalamic stroke, indicating that lesion location (e.g., basal ganglia vs. thalamus) and hemispheric laterality (e.g., left vs. right hemisphere) also play a role in the degree of language impairment.

There are other potential neural causes of subcortical aphasia. The co-occurrence of subcortical stroke and symptoms of small vessel disease and prior subcortical ischemia is high (Paradise et al., 2018). In particular, leukoaraiosis is a form of white matter disease characterized by white matter dysfunction due to perfusion disturbances within arterioles that perforate the deep brain structures (Marek et al., 2018). These diffuse hyperintensities are associated with an increased risk of cognitive impairment (Te et al., 2015; Yuan et al., 2018), and their prevalence increases with age, especially after age 60 (Marek et al., 2018; Vedala et al., 2019; Wright et al., 2018). Leukoaraiosis is more prevalent in males (Henninger et al., 2013) and has been linked to poor outcomes in acute cortical stroke (Arba et al., 2016; Fierini et al., 2017), including a 4.3-fold increase in chronic decline of language abilities (Basilakos et al., 2019). In addition, leukoaraiosis severity is predictive of infarct volume (Henninger et al., 2014), infarct growth (Ay Hakan et al., 2008), and response to language therapy in cortical stroke (Varkanitsa et al., 2020). Whether these trends exist in subcortical stroke populations is unclear.

The present study aimed to disentangle the independent contributions of each of these factors (i.e., lesion volume and location, hypoperfusion, leukoaraiosis, prior infarcts) to language impairments in acute (less than 10 days post-stroke onset) left hemisphere subcortical stroke. We hypothesized that greater hypoperfusion, more severe leukoaraiosis, and higher total lesion volume (including damaged tissue from prior subcortical stroke) will serve as strong negative predictors of acute language abilities. As a precursor to our main aim, we also determined relationships between stroke factors and demographic variables such as age and sex.

The importance of determining which of these factors contribute to subcortical aphasia is twofold. First, characterizing subcortical participation in language functions (e.g., lexical-semantic processing), if any, will deepen our understanding of these structures beyond their well-characterized roles in relaying sensory and motoric cortical inputs and outputs. Second, the present study may guide clinicians’ decisions to look beyond infarct volume for predictors of acute subcortical aphasia. That is, if leukoaraiosis, hypoperfusion, and/or a history of prior stroke can independently account for the incidence of subcortical aphasia, those findings may form a more complete clinical picture of subcortical stroke during its arguably most critical window.

## 2. Methods

### 2.1. Participants

We retrospectively reviewed 1043 records of individuals who were admitted to Johns Hopkins Hospital or Bayview Medical Center between 2002 and 2019 and recruited as part of a larger study aimed at investigating left hemisphere stroke recovery. The final sample included 80 individuals (36 women, mean age = 55.7 ± 14.5 years) who were confirmed to have an acute LH subcortical stroke *without* cortical involvement or acute lesion elsewhere in the brain (e.g., brainstem, cerebellum). Our final sample comprised 66 individuals (29 women, mean age = 55.7 ± 14.5 years) who had sufficient language data for analysis. A subset of our sample (n=28) had prior strokes restricted to subcortical structures in one or both hemispheres (left=6, right=8, bilateral=14) in addition to their acute LH subcortical lesion. Patients with underlying neuropathology (e.g., Alzheimer’s disease) besides stroke were excluded so that any language impairments present could be attributed to stroke and not to other causes. Ethics approval was obtained from the Johns Hopkins University institutional review board, and written informed consent was provided by all participants prior to enrolling in the study.

### 2.2. Neuroimaging Data

Upon their acute hospitalization, each patient underwent a clinical magnetic resonance imaging (MRI) protocol that included diffusion-weighted (DWI), T2-weighted, and fluid-attenuated inversion recovery (FLAIR) sequences. The average time interval between the acute stroke and clinical imaging acquisition was 0.89 ± 1.07 days.

Using MRIcron (Rorden & Brett, 2000), trained technicians manually delineated acute lesions slice-by-slice on DWI scans, which are highly sensitive to stroke lesions in the acute phase (Hillis et al., 2002). Similarly, chronic lesions were manually delineated on T2-weighted imaging for the 28 individuals with prior subcortical involvement. From these tracings, acute lesion volume, total lesion volume (acute + chronic volumes for participants with prior stroke), and the percentage of damaged tissue in subcortical regions of interest (ROIs) were calculated using NiiStat (http://www.nitrc.org/projects/niistat). Subcortical ROIs included the basal ganglia (caudate nucleus, putamen, globus pallidus), thalamus, superior and posterior corona radiata, anterior and posterior limbs of the internal capsule, and external capsule. See **Figure 1A** for a visualization of acute lesions on DWI for three sample participants and **Figure 1B** for visualizations of prior lesions.

**Fig. 1.**
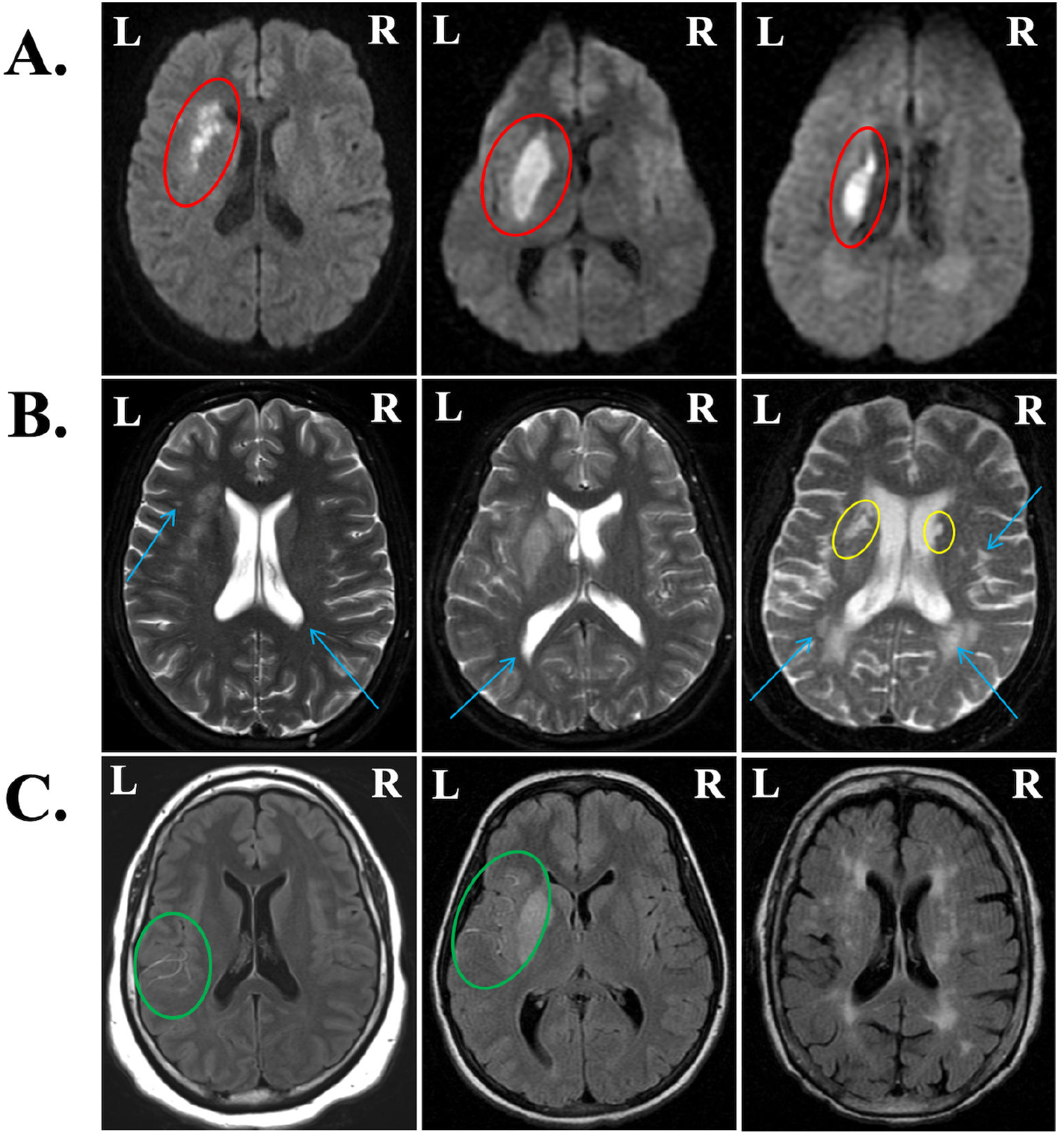
Representative images from patients with varying acute and chronic infarct volumes, leukoaraiosis, and hypoperfusion. (A) Diffusion-weighted images (DWI) display acute lesions (red ovals). (B) T2-weighted scans were used to visually score leukoaraiosis on the Fazekas scale (blue arrows) from 0 to 3 and visualize chronic infarcts where present (yellow ovals). From left to right, periventricular hyperintensities (PVH) were scored as 1, 1, and 3; deep white matter hyperintensities (DWMH) were scored as 1, 0, and 2. (C) FLAIR hyperintense vessels (FHV) are indicative of hypoperfusion in areas surrounding infarcted tissue (green ovals). From left to right, FHV scores were 5, 7, and 0 out of 12.

The severity of leukoaraiosis both ipsi- and contralateral to the acute LH infarct was visually assessed on T2-weighted MRI imaging and rated using the Fazekas scale (Fazekas et al., 1987), which comprises two qualitative subscales. The first captures periventricular hyperintensities (PVH), found near the horns of the lateral ventricles, and is scored from 0 (no hyperintensities) to 3 (irregular hyperintensities extending into the deep white matter). The second subscale evaluates the severity of deep white matter hyperintensities (DWMH) and is also scored from 0 (no hyperintensities) to 3 (large confluent hyperintense areas). PVH and DWMH subscale ratings were made independently for the left and right hemisphere by two independent, trained raters (M.S. and E.M.). Consensus ratings were given when ratings by M.S. and E.M. differed by more than one point. Because chronic stroke lesions also appear hyperintense on T2-weighted images, the manual tracing of the prior stroke lesion was overlaid on the T2-weighted image while the leukoaraiosis ratings were completed for the 28 individuals with a history of subcortical stroke recurrence. The T2-weighted image and associated leukoaraiosis ratings for three sample patients are shown in **Figure 1B**.

To prepare the Fazekas data for analysis, we next compared left and right hemisphere (RH) Fazekas ratings using Wilcoxon rank sum tests. Leukoaraiosis is often symmetrical between hemispheres, and indeed, we found no significant hemispheric differences for either PVH (W = 3307, p = 0.697) or DWMH (W = 3063, p = 0.617) ratings. In contrast, within each hemisphere, PVH ratings significantly differed from DWMH ratings (LH: W = 3943, p = 0.007; RH: 3977, p = 0.004). Therefore, we averaged the ratings from each hemisphere to generate a single PVH and single DMWH rating for each participant. Inter-rater reliability was calculated according to Cohen’s weighted kappa (κ_w_ = 0.676, p < 0.001 for PVH scores; κ_w_ = 0.457, p < 0.001 for DWMH scores).

Regions of hypoperfusion often extend beyond the lesioned tissue itself, which offers additional information about the extent of possible language deficits (Payabvash et al., 2011). In clinical sequences, perfusion-weighted imaging (PWI) is often used to quantify hypoperfusion, yet the limited availability of perfusion data in our sample made a PWI approach to assessing hypoperfusion untenable. Notably, arteries supplying hypoperfused areas of the brain appear bright (hyperintense) on FLAIR imaging due to slow blood flow (Reyes Dennys et al., 2017). Therefore, we evaluated FLAIR scans for hyperintense vessels in each of the vascular territories that supply the brain with perfused blood. Reyes Dennys et al. (2017) found that FLAIR hyperintense vessels (FHV) are a reliable surrogate for PWI data and accurately reflect the burden of hypoperfusion in the acute phase. We employed the NIH-FHV scoring system used by Reyes et al., which rates the number of hyperintense vessels per axial slice in each of six vascular territories in the affected (left) hemisphere. The vascular territories assessed were those of the anterior cerebral artery, posterior cerebral artery, and the frontal, temporal, parietal, and insular divisions of the middle cerebral artery. In each territory, a score of 0 denotes no FHVs in any slice and a score of 2 denotes ≥ 3 FHVs per slice or FHVs present in ≥ 3 axial slices. To index overall hypoperfusion, we summed all territory FHV ratings to generate a single total FHV measure. Total FHV scores for each patient (78/80 received FLAIR imaging) could range from 0 to 12. See **Figure 1C** for a visualization of FHVs in three example participants. Inter-rater reliability for the total FHV rating between the two trained raters (M.S. and A.W.) was calculated according to Cohen’s weighted kappa (κ_w_ = 0.934, p < 0.001). Notably, the distribution of total FHV scores was heavily skewed to the lower end of the scale, with 47 participants with no FHVs (i.e., scores of 0). As such, the FHV rating was transformed into a dichotomous variable such that 0 reflected no hypoperfusion (i.e., 0 FHVs) and 1 reflected some degree of hypoperfusion (i.e., ≥ 1 FHV).

### 2.3. Language Assessments

Because our sample draws from patient data that spans 17 years (2002-2019), participants were administered a variety of language assessments. Participants received either the Boston Diagnostic Aphasia Examination (BDAE, n=15) (Goodglass et al., 2001a), Western Aphasia Battery-Revised (WAB-R, n=25) (Kertesz et al., 2007), or an in-house lexical battery (LB, n=30). Language data from 14 of the original 80 included patients could not be located or were incomplete. The average time interval between the stroke date and administration of the language battery was 1.56 ± 1.50 days, while the average gap between clinical imaging acquisition and language battery administration was 0.84 ± 1.27 days.

These batteries, while assessing similar components of language ability, employ unique tasks and scoring systems. Equivalent subtests were extracted from each battery to index auditory comprehension, naming, and verbal expression skills (see Supplemental Table 1). To approximate equivalent measures across the three batteries, a binary aphasic variable was generated based on metrics particular to each battery. Kertesz et al. (2007) established an aphasia quotient (AQ) of 93.8 or less (out of 100) to denote aphasic status on the WAB-R, while Hillis et al. (2002) demarcated a score of 89% or below on the LB to denote aphasic status. No published cutoff for aphasic/non-aphasic status is readily available for the BDAE, so z-scores were calculated within the subset of recipients of each battery. The z-scores corresponding to the published aphasia cutoffs on the WAB-R and LB were similar (WAB-R z-score = -0.308, LB z-score = -0.336). Thus, the average of the z-scores corresponding to the LB and WAB cutoffs was used to establish the aphasic/non-aphasic cutoff for the BDAE (z = -0.322).

### 2.4. Statistical Analysis

Brain predictors of interest included: total lesion volume; a binary stroke history variable (i.e., 0 = no history of prior stroke, 1 = history of prior stroke) to capture stroke recurrence; percent damage to ROIs (i.e., thalamus, basal ganglia, corona radiata, internal capsule, and external capsule) to index lesion location; the binary FHV measure to capture hypoperfusion extent; and PVH and DWMH ratings within each hemisphere to measure leukoaraiosis. First, to determine relationships between brain variables and demographics, we conducted Wilcoxon rank sum or chi-square tests to determine if categorical variables (i.e., stroke history, hypoperfusion, and leukoaraiosis measures) varied between sexes and Spearman correlations to examine relationships between continuous variables (i.e., lesion volume, percent damage to ROIs) and age. We corrected for multiple comparisons at a false discovery rate (FDR) of q > 0.05. We also compared participants with and without aphasia in age and sex using Wilcoxon rank sum and chi-square tests, respectively. To address our main aim, we compared brain variables of interest between participants with and without aphasia using Wilcoxon rank sum and chi-square tests, correcting for multiple comparisons using the FDR correction within each metric type (i.e., damage, leukoaraiosis, or hypoperfusion). Finally, to determine the relative contributions of each variable on presence of aphasia, we conducted a logistic regression including all univariate predictors that significantly differed between people with and without aphasia before correction and aphasia status (i.e., aphasic/not aphasic) as the dependent variable.

## 3. Results

### 3.1. Demographic Relationships

While sex and leukoaraiosis were not significantly associated (PVH: p = 0.565, q = 0.633; DWMH: p = 0.633, q = 0.633), older age *was* linked to more severe leukoaraiosis (PVH, DWMH: p < 0.001, q < 0.001). Binary hypoperfusion status in accordance with FHV ratings was not linked to either age (p = 0.537, q = 0.537) or sex (p = 0.348, q = 0.537). No other significant relationships between demographic and stroke variables were found. Aphasia status did not differ between men and women (p = 0.7103), but participants with aphasia were significantly older than participants without aphasia (p = 0.021).

### 3.2. Relationships between Brain Variables and Aphasia Status

Participants without FLAIR scans and/or with missing or incomplete language data were excluded from the analyses aimed at investigating how brain metric varied by aphasia status. A subset of 66 patients was included in the final analyses. Of these, 21 patients were designated as aphasic (31.8%) according to the aforementioned cutoffs, and 45 were not aphasic.

Compared to individuals without aphasia, those with aphasia had larger acute lesion volumes (p = 0.033, q = 0.033) as well as larger *total* lesion volumes (p = 0.012, q = 0.025), which included both the acute LH and prior subcortical lesions. Yet, the number of participants with a history of prior stroke did not differ between groups (p = 0.155). An overlap map for both acute and chronic lesions in our sample is shown in **Figure 2**. Patients with aphasia had greater percent damage to the basal ganglia (p = 0.041, q = 0.069), corona radiata (p = 0.031, q = 0.069), and internal capsule (p = 0.003, q = 0.014) than did patients without aphasia, but only the internal capsule result survived a correction for multiple comparisons. Ultimately, none of these findings survived a correction for stroke volume (p ≥ 0.168). We found no significant differences in leukoaraiosis between aphasic/non-aphasic groups by either Fazekas subscale (PVH: p = 0.088, q = 0.175 ; DWMH: p = 0.255, q = 0.255). Similarly, presence of hypoperfusion did not significantly differ between patients with versus without aphasia. See **Table 1** for a summary of all brain data by group.

**Fig. 2.**
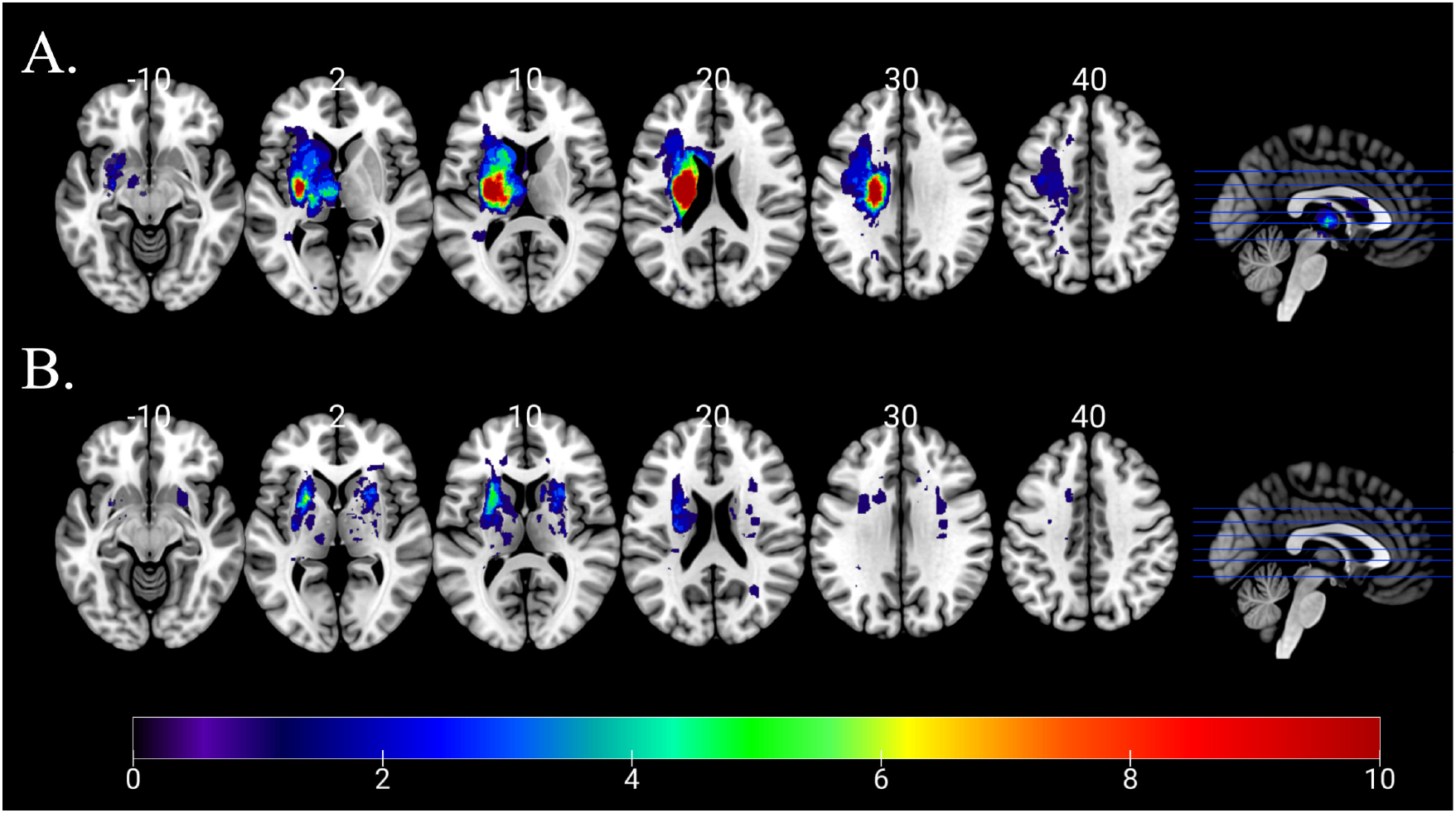
Lesion overlap maps of A) acute stroke lesions and B) prior stroke lesions. The color bar applies to both parts of the figure and indicates the number of patients whose lesions overlap for a given voxel.

**Table 1.**
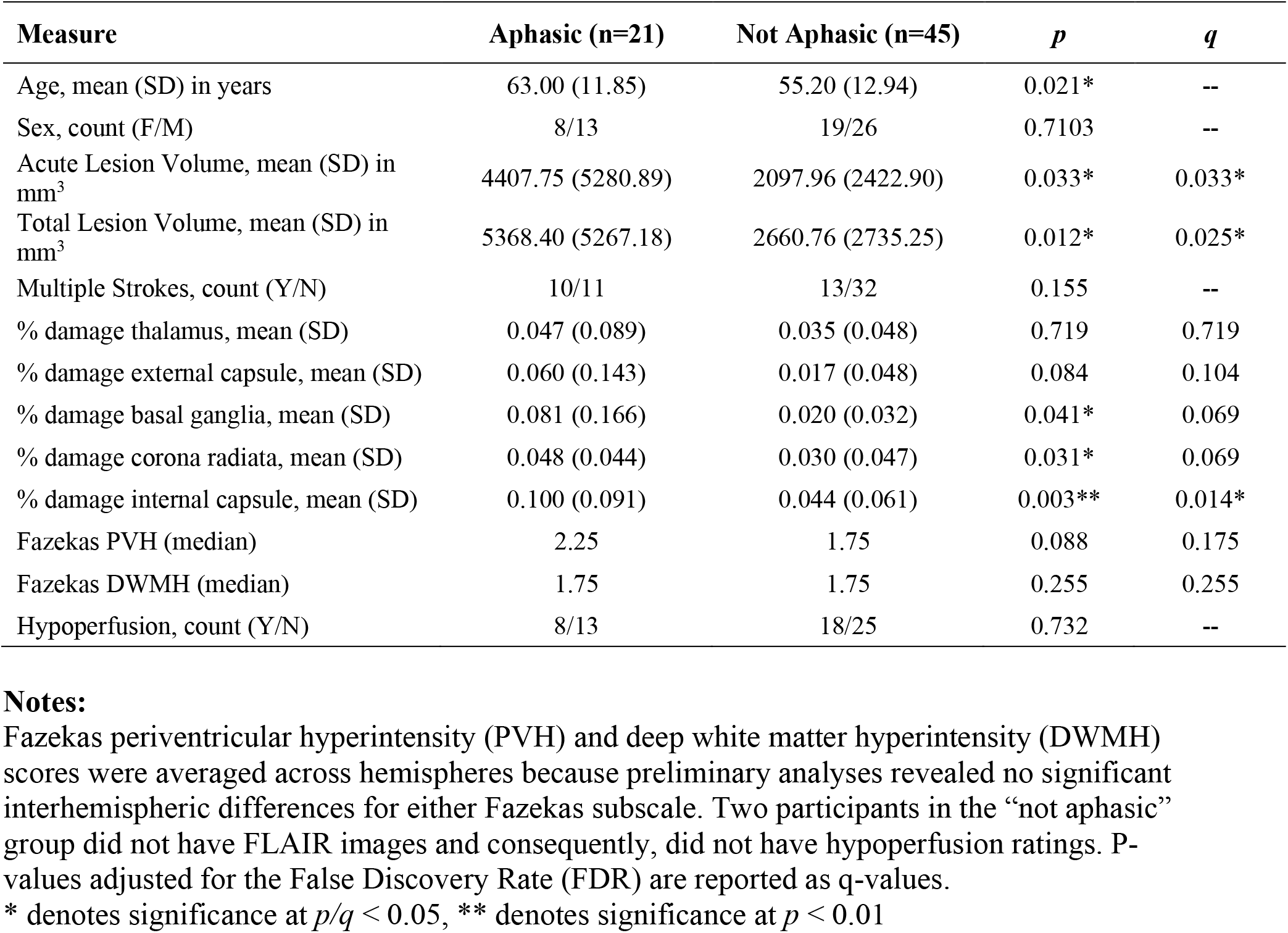
Demographic and stroke variables compared between aphasic and non-aphasic participants.

| Measure | Aphasic (n=21) | Not Aphasic (n=45) | *p* | *q* |
| --- | --- | --- | --- | --- |
| Age, mean (SD) in years | 63.00 (11.85) | 55.20 (12.94) | 0.021* | -- |
| Sex, count (F/M) | 8/13 | 19/26 | 0.7103 | -- |
| Acute Lesion Volume, mean (SD) in mm<sup>3</sup> | 4407.75 (5280.89) | 2097.96 (2422.90) | 0.033* | 0.033* |
| Total Lesion Volume, mean (SD) in mm<sup>3</sup> | 5368.40 (5267.18) | 2660.76 (2735.25) | 0.012* | 0.025* |
| Multiple Strokes, count (Y/N) | 10/11 | 13/32 | 0.155 | -- |
| % damage thalamus, mean (SD) | 0.047 (0.089) | 0.035 (0.048) | 0.719 | 0.719 |
| % damage external capsule, mean (SD) | 0.060 (0.143) | 0.017 (0.048) | 0.084 | 0.104 |
| % damage basal ganglia, mean (SD) | 0.081 (0.166) | 0.020 (0.032) | 0.041* | 0.069 |
| % damage corona radiata, mean (SD) | 0.048 (0.044) | 0.030 (0.047) | 0.031* | 0.069 |
| % damage internal capsule, mean (SD) | 0.100 (0.091) | 0.044 (0.061) | 0.003** | 0.014* |
| Fazekas PVH (median) | 2.25 | 1.75 | 0.088 | 0.175 |
| Fazekas DWMH (median) | 1.75 | 1.75 | 0.255 | 0.255 |
| Hypoperfusion, count (Y/N) | 8/13 | 18/25 | 0.732 | -- |

**Notes:**
Fazekas periventricular hyperintensity (PVH) and deep white matter hyperintensity (DWMH) scores were averaged across hemispheres because preliminary analyses revealed no significant interhemispheric differences for either Fazekas subscale. Two participants in the “not aphasic” group did not have FLAIR images and consequently, did not have hypoperfusion ratings. P-values adjusted for the False Discovery Rate (FDR) are reported as q-values.
\* denotes significance at $p/q < 0.05$ , \*\* denotes significance at $p < 0.01$

### 3.3. Logistic Regression Predicting Aphasia Status from Significant Univariate Predictors

Last, we conducted a logistic regression that included all univariate predictors that differed significantly between people with and without aphasia (before correction), including age, total lesion volume, and percent damage to the basal ganglia, internal capsule, and corona radiata. Within this model, only age significantly predicted aphasia status (p = 0.014). See **Table 2** for the full model results.

**Table 2.** Logistic regression predicting aphasia status from significant univariate predictors.

| Variable | Estimate | Standard Error | z score | p-value |
| --- | --- | --- | --- | --- |
| Intercept | -6.486 | 2.067 | -3.138 | 0.002 |
| Basal Ganglia ROI | -3.459 | 7.100 | -0.487 | 0.626 |
| Corona Radiata ROI | -1.625 | 7.276 | -0.233 | 0.823 |
| Internal Capsule ROI | 6.608 | 6.232 | 1.060 | 0.289 |
| Total Lesion Volume | 0.000 | 0.000 | 1.689 | 0.091 |
| Age | 0.071 | 0.029 | 2.450 | 0.014 |
| Days Between Stroke and Test | 0.154 | 0.200 | 0.679 | 0.442 |

Residual deviance: 64.766 on 59 degrees of freedom. Null deviance: 80.970 on 65 degrees of freedom. Akaike Information Criterion (AIC): 78.766.

While subcortical stroke can cause aphasia, it usually does not — and when it does, the deficits tend to be mild. Despite having 21 individuals who were classified as aphasic, most had z-scores that were close to their respective cutoff and presented with mild deficits. The significant differences before correction were driven by three patients in particular with marked language deficits (z-scores 1.5 or more standard deviations below the sample mean). Their imaging and language assessment profiles were examined further.

### 3.4. Patients with Severe Language Deficits

#### 3.4.1. Patient 1

Patient 1 was an early 70’s man with no prior strokes who presented with total anarthria (absence of speech). This patient used a communication board and was able to nod or shake his head. He received the WAB-R and performed 2.7 standard deviations below the mean (AQ=55.4). Patient 1’s auditory comprehension for simple yes/no questions was intact, and single-word auditory comprehension was also mostly intact (54/60). Meanwhile, sentence comprehension for complex syntactic structures, notably those of multistep commands, was impaired. The patient’s forward word span was mildly impaired, but backwards span was severely impaired. Patient 1’s DWI revealed a large LH basal ganglia lesion (corpus striatum, 20,090 mm^3^) that extended into the periventricular white matter. Periventricular leukoaraiosis was rated 2/3 and DWMH was rated 1/3. Patient 1 was given an FHV score of 1/12 as a measure of hypoperfusion. See **Figure 3A** for a visualization of Patient 1’s imaging data.

**Fig. 3.**
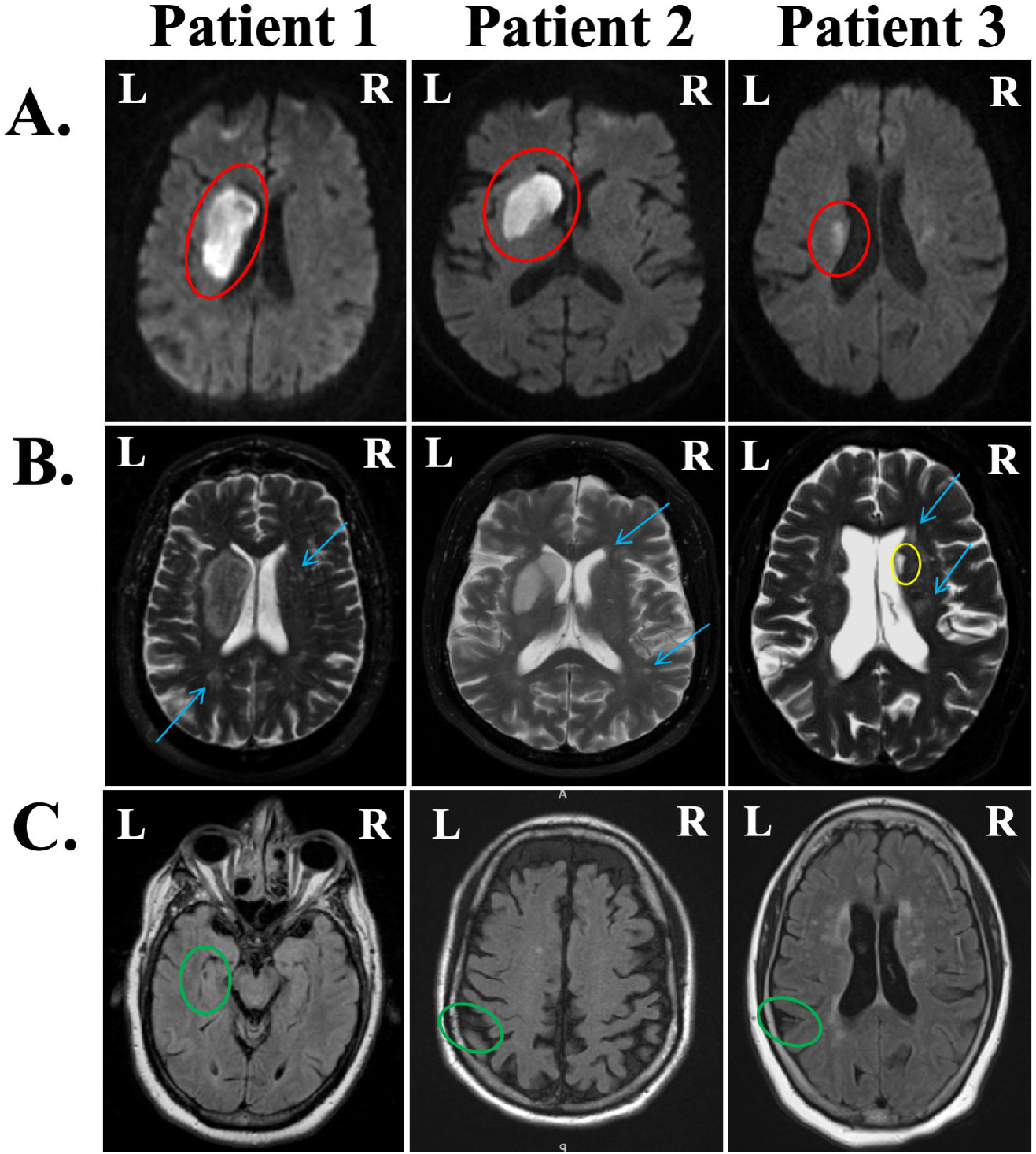
Clinical images from the three patients with marked language deficits in the acute phase. (A) Diffusion-weighted images (DWI) display acute lesions (red ovals). Patients 1 and 2 had lesions restricted to the basal ganglia, and Patient 3 had lesions to the basal ganglia and internal capsule. (B) T2-weighted scans were used to visually score leukoaraiosis on the Fazekas scale (blue arrows) from 0 to 3 and visualize chronic infarcts where present (yellow ovals). From left to right, periventricular hyperintensities (PVH) were scored as 2, 1, and 2; deep white matter hyperintensities (DWMH) were scored as 1, 1, and 2. (C) FLAIR hyperintense vessels (FHV) are indicative of hypoperfusion in areas surrounding infarcted tissue (green ovals). Only Patient 3 had a prior stroke. All three patients received FHV scores of 1 out of 12.

#### 3.4.2. Patient 2

Patient 2 was a mid-60’s man with no prior strokes who presented with severely impaired word fluency. For example, when asked to name as many animals as possible, Patient 2 said “Pork and beans, camel, bacon, pork, a whole bunch of animals that eat pork.” The patient was hospitalized for nearly a month after his stroke due to a fall with possible loss of consciousness (which co-occurred with the stroke). He received the WAB-R and performed 3.055 standard deviations below the mean (AQ=76.9). Patient 2 made unique word errors in expressive naming and sentence completion tasks; he used off-target, unrelated words, and also produced some semantic paraphasias (e.g., using *envelope* for *mail, May* for *December*). As with Patient 1, Patient 2 was impaired with multistep commands (55/80). This patient’s stroke was localized to the left basal ganglia (caudate and lentiform nuclei, 16.871 mm^3^). Both PVH and DWMH scores were 1/3, and hypoperfusion was rated 1/12. See **Figure 3B** for a visualization of Patient 2’s imaging data.

#### 3.4.3. Patient 3

Finally, Patient 3 was a late 50’s woman admitted to Johns Hopkins Hospital for previous symptomatic infarcts, presenting with dysarthria but not aphasia. Patient 3 had bilateral basal ganglia involvement (right striatocapsular region and both caudate heads). She presented with left-sided weakness, and received the BDAE, performing 2.2 standard deviations below the mean. This patient’s auditory comprehension was intact (15/16), and she was able to follow multistep commands well. Patient 3 was able to produce some single words, but rarely surpassed one-word phrase lengths (1/7 grammatical form, 2/7 articulatory agility on the Rating Scale Profile of Speech Characteristics within the BDAE). In addition, Patient 3’s sentence repetition was poor (0-20^th^ percentile). Patient 3’s DWI revealed an acute LH infarct localized to the basal ganglia and internal capsule (1523 mm^3^). The patient’s prior bilateral subcortical lesions totaled 776 mm^3^, for a total lesion volume of 2299 mm^3^. Both PVH and DWMH scores were 2/3, so perhaps the severity of this patient’s leukoaraiosis contributed to her deficits. The patient’s total hypoperfusion was rated 1/12. See **Figure 3C** for a visualization of Patient 3’s imaging data. Comprehensive language data for all three patients are reported in Supplemental Table 2.

## 4. Discussion

In this study, we investigated the contribution of multiple brain metrics on the presence of aphasia in acute left hemisphere subcortical stroke. Overall, we found differences between patients with versus without aphasia in age and lesion volume (and to a lesser extent location, prior to correction for multiple tests). The majority null findings are important from a clinical and scientific standpoint in this population. Below, we address each finding in greater detail and situate the results in the context of previous literature.

### 4.1. The Frequency and Severity of Acute Subcortical Aphasia

In our study, we found that 68.2% of patients were not aphasic, whereas only 31.8% were aphasic. The prevalence of aphasia in acute subcortical stroke has varied widely. Hillis et al. (2002) reported aphasia in 25 out of 37 patients (67.6%) with acute strokes restricted to subcortical structures. In a later study, Hillis et al. (2004) reported an aphasia frequency of 54.1% in a patient corpus of 24 patients, although 16 of these patients were also included in the earlier study. Interestingly, another study by Choi et al. (2007) found that 15 of 16 patients (93.8%), all of whom had acute left striatocapsular strokes and were administered the Korean version of the WAB, were aphasic. Olsen et al. (1986), on the other hand, found that only 8 of 18 patients (44.4%) with acute subcortical LH lesions were aphasic. That the majority of patients in the study by Olsen et al. (1986) did not even exhibit minimal speech-language deficits to meet the criteria for mild aphasia suggests a high proportion of acute LH subcortical stroke patients whose language is completely intact.

One key difference across these studies, including our own, is the use of different language batteries and scales to measure aphasia. Both studies by Hillis et al. (2002, 2004) used the lexical battery, and the later study also used the BDAE for measures of repetition, auditory comprehension, and naming. Choi et al. (2007) instead used the WAB, which measures spontaneous speech generation, auditory comprehension, repetition, and naming. Our study used data from an in-house lexical battery, BDAE, and the WAB, and extracted measures of auditory comprehension, naming, and verbal expression skills. Meanwhile, Olsen et al. (1986) used an aphasia severity scale proposed by Goodglass et al. (2001b), which assigns a score from 0 to 3 based on qualitative language outcomes such as “conversation about familiar subjects is possible with help from the listener” (score 2). In addition to testing different domains of language, the different batteries may have varying sensitivities to aphasia. Of note in our study, however, is the largest patient sample, which is perhaps more representative of the acute LH subcortical stroke population than earlier studies.

Accounts of the severity of LH subcortical aphasia are similarly inconsistent. Only three patients in our sample (4.5%) presented with severe language deficits (language z-scores more than 1.5 standard deviations below the mean). Many subjects who *were* aphasic bordered established cutoffs for the binary aphasic variable in their respective language batteries. For example, six of eight aphasic patients who received the WAB scored above 90 but below the aphasia cutoff of 93.8. Of the eight aphasic patients in the study by Olsen et al. (1986), only one had severe aphasia, whereas three had mild aphasia and the last three had moderate aphasia. We are led to believe, then, that when language deficits do occur after LH subcortical stroke, they are not often severe. While the distributions of severe versus borderline cases of aphasia in the other aforementioned studies are not reported, the authors of the later study by Hillis et al. (2004) acknowledge inconsistencies in aphasia severity due to subcortical infarcts.

### 4.2. Relationship Between Lesion Location and Aphasia Status

#### 4.2.1. Thalamus

The link between overall brain damage, captured by total lesion volume, and deficit severity is well-established. The relationship between lesion site and incident aphasia, however, is more debated. Numerous functional imaging and clinical studies have identified thalamic involvement in language processing (Bogousslavsky & Caplan, 1993; Crosson, 1984, 1985; Llano, 2013; Nadeau & Crosson, 1997) — and left thalamic damage has previously been linked to language deficits and the presence of aphasia, but not in the present study. That is, our results do not corroborate the link between thalamic damage and language deficits found in prior studies.

Accounts in the literature are, however, mixed. Hillis et al. (2002), for example, found that 0 of 5 patients with isolated LH thalamic strokes were aphasic. On the other hand, Karussis et al. (2000) reported aphasia in 88% (n=8) of acute left thalamic stroke patients, and a meta-analysis by De Witte et al. (2011) reported aphasia in 64% (n=11) of patients with acute left thalamic lesions, where the label of thalamic aphasia required deficits in at least four of the following six categories: fluency, comprehension, repetition, naming, reading, and writing. Further, Sebastian et al. (2014) found that among a sample of 10 patients with isolated left hemisphere thalamic stroke (six men, four women, mean age 45.3 ± 8.8 years), half were aphasic. Of the five aphasic patients, four had normal cortical perfusion. While we, too, found that subcortical aphasia can occur without cortical hypoperfusion, we did not find a relationship between any region of interest (i.e., left thalamus) and presence of aphasia that survived a correction for multiple comparisons and control for total lesion volume. Recently, a study by Rangus et al. (2021) found that 48% (n=31) of patients with isolated acute LH thalamic lesions met the diagnostic criteria for aphasia. The discordance between our study and those proposing a link between thalamic aphasia and language deficits might be due to our sample’s low proportion of thalamic stroke patients, who themselves did not present with large thalamic lesions. Only 13 patients out of 80 (16.4%) had greater than 10% damage to the left thalamus (range = 0-31.6% damage). Of these 13 patients with significant thalamic damage, only four were aphasic (30.7%). Moreover, of the 21 aphasic patients in our sample, only four patients (19%) had greater than 10% damage to the left thalamus. Overall, then, the frequency of thalamic lesions among purely subcortical lesions was low in our study, as was the frequency of aphasia among patients with significant thalamic damage.

The literature also contains discrepant reports of the symptoms of individuals with thalamic aphasia. The aforementioned study by De Witte et al. (2011) found, for example, that 14 of 32 (43.8%) patients with acute LH thalamic lesions presented with language comprehension problems (although only 3 of these cases amounted to “severe comprehension disturbances”) and only one patient presented with non-fluent speech. Strikingly, the authors report that of 26 patients with LH thalamic lesions, 18 had moderate to severe naming problems. The comparability of this analysis to the present study is limited by the use of different diagnostic criteria for thalamic aphasia (i.e., the inclusion of repetition, reading, and writing data). In the aforementioned study by Rangus et al. (2021), the authors found that when present, language deficits secondary to LH thalamic lesions were mild; 67% of patients exhibited either a letter fluency or semantic fluency deficit, but 96% showed fluent, spontaneous speech. Moreover, naming deficits were only found in 19% of patients with LH thalamic lesions, and auditory and verbal comprehension were intact. This account is largely consistent with our finding that when thalamic lesions do occur, they rarely lead to severe language deficits. That naming and auditory comprehension were largely unaffected in the study by Rangus et al. justifies the results of our analysis because the binary aphasic variable in our study relied upon metrics of naming, auditory/verbal comprehension, and verbal expression.

The nucleus of the thalamus that is infarcted is almost certainly important in determining whether or not aphasia is present. The various nuclei of the thalamus are known to be involved in distinct functions (e.g. attention, somatosensory function, motor relay, limbic functions). However, it is difficult to localize the specific nucleus of the thalamus on clinical MRI. Doing so remains an important future direction for future research involving high resolution research MRI in which segmentation of thalamic nuclei is feasible.

#### 4.2.2. Basal Ganglia

As with the thalamus, there is little consensus surrounding the role of the basal ganglia in language. In our sample, patients with aphasia had greater percent damage to the basal ganglia, but this finding did not remain significant after correcting for multiple comparisons or controlling for lesion volume. A recent meta-analysis by Radanovic & Mansur (2017) illuminates the state of the literature in characterizing the frequency and phenotype of basal ganglia stroke. Their review generated 180 patients from 57 separate studies with acute left hemisphere lesions restricted to the basal ganglia, with or without damage to the internal capsule. Overall, 67.3% had language deficits of some sort, although this incidence varied greatly depending on the structure affected. For example, patients whose damage was isolated to the caudate (n=24), putamen (n=31), or globus pallidus (n=4) had language disturbances at a rate of 41.7%, 80.7%, and 0%, respectively. Patients with isolated caudate or putamen damage had heterogeneous language symptoms with few associations drawn. The production of paraphasias was linked to damage to the putamen (odds ratio (OR) of 2.71), whereas repetition impairments and nonfluent aphasia were less associated with caudate lesions (OR of 0.17 and 0.28, respectively).

Within our sample of 80 patients, significant damage to structures within the basal ganglia was uncommon. When such damage *did* occur, it was rarely isolated to a single structure, and rarely was it exclusive of the surrounding white matter. Few patients had greater than ten percent damage to the caudate nucleus (10.0%), putamen (11.3%), and globus pallidus (8.8%). Of the eight patients with significant damage to the left caudate, only two had lesions that did not extend to the globus pallidus or putamen, and none had lesions exclusive of the internal capsule. Similarly, of the nine patients with significant putaminal damage, only three had lesions exclusive of the caudate and globus pallidus, and all of their lesions extended into the internal capsule. Finally, of the seven patients with significant pallidal involvement, only three spared the caudate or putamen, and only one of those three had a lesion that did not extend to the surrounding white matter. Clearly, our sample population did not lend itself to an analysis that isolated particular basal ganglia to examine their roles in LH subcortical aphasia. Had our sample contained more patients with significant and exclusive basal ganglia involvement, it is not clear that our results would change, as our language data lacked measures of repetition — one of the few components of language associated with lesions isolated to single structure.

When multiple basal ganglia are affected, with or without involvement of the adjacent white matter, accounting for the clinical picture of these patients becomes more difficult. Let us return to the meta-analysis by Radanovic & Mansur. Taken alone, lesions to the putamen were only associated with paraphasia, and pallidal lesions were not associated with any language symptoms. However, in patients with strokes isolated to the lentiform nucleus (which comprises the putamen and globus pallidus), the authors report a higher likelihood of repetition impairment, comprehension deficit, and nonfluent aphasia (OR of 5.78, 3.50, 3.23, respectively). Interestingly, lesions to the lentiform nucleus were also associated with the *absence* of language disturbances (OR of 2.65) (Radanovic & Mansur, 2017). Further, the authors found that neither striatal (affecting the caudate and putamen) nor striatopallidal lesions (affecting the caudate, putamen, and globus pallidus) were associated with language symptoms. When white matter involvement was considered, the constellation of language symptoms remained heterogeneous. Patients with striatocapsular infarcts, for example, presented with anomia (47%), repetition deficits (35.3%), comprehension impairments (32.3%), and paraphasias (32.3%). The authors do note, however, that the extent of white matter damage plays a role in determining the severity of nonfluency (Radanovic & Mansur, 2017).

For the most part, taking larger subsets of basal ganglia structures, with or without inclusion of the surrounding white matter, does not reveal decisive clinicoanatomical correlations with language symptoms. In those cases where particular subsets of structures *do* reveal characteristic language symptoms, the present study was unable to capture those relationships, due in large part to the scarcity of patients with significant basal ganglia involvement and the lack of repetition data.

### 4.3. Leukoaraiosis

In our study, we found that more extensive leukoaraiosis severity was related to older age but not the presence of aphasia after subcortical stroke. We did not corroborate findings that leukoaraiosis is more prevalent in men (Henninger et al., 2013). The finding that greater leukoaraiosis severity is linked with greater age has considerable support in the literature. The Leukoaraiosis and Disability (LADIS) study examined the major determinants of leukoaraiosis severity and found that age, frequency of hypertension, and prior stroke history increased with leukoaraiosis severity as measured by the Fazekas scale (Basile et al., 2006). This study of 639 patients included nondisabled elderly subjects and did not focus on stroke patients specifically.

In a study of neglect performance in acute right hemisphere stroke, Bahrainwala et al. (2014) reported significantly higher age among patients (n=205) with severe leukoaraiosis compared to patients with milder leukaraiosis, as measured by the Cardiovascular Health Study (CHS) rating scale from 0-9 (Manolio et al., 1994). The authors did not separate cortical stroke from subcortical stroke patients, although they did not expect that doing so would alter their findings (Bahrainwala et al., 2014). More recently, a meta-analysis by Vedala et al. (2019) found using the Wahlund leukoaraiosis scale, which ranges from 0 to 30 and accounts for hemispheric differences as well as regional differences, that leukoaraiosis was most severe in older patients (Wahlund et al., 2001). The authors found this to be true on both MRI (n=186) and CT (n=238) clinical imaging. Importantly, our concordance with previous studies on the link between age and leukoaraiosis severity validates our chosen method of measuring leukoaraiosis.

Prior studies have shown relationships between leukoaraiosis extent and the severity of post-stroke deficits in domains outside of language. For example, Arsava et al. (2009) manually delineated white matter lesions and found a moderate correlation between leukoaraiosis volume and clinical outcomes at 6 months post-stroke as measured by the modified Rankin Scale (mRS), which reflects neurologic disability after stroke but not language in particular. Henninger et al. (2013) and Arba et al. (2016) had similar findings for 90-day outcomes, also using the mRS. A review by Fierini et al. (2017) found that greater leukoaraiosis severity predicts poorer overall clinical outcomes in the acute and subacute phases.

From the acute to chronic stages of cortical stroke recovery, it appears that greater leukoaraiosis is associated with poorer neurological outcomes. Such a relationship is not so clear for language in particular. A few studies have investigated the relationship between leukoaraiosis and language deficits in patients with aphasia due to cortical stroke. Wright et al. (2018) examined language outcomes and found that in patients greater than three months post-stroke (well outside the acute window), leukoaraiosis severity as measured by the CHS scale was negatively correlated with object naming and word fluency performance after controlling for lesion volume. A retrospective study by Varkanitsa et al. (2020) aimed to identify how leukoaraiosis factors into pre- and post-treatment language outcomes in 30 patients with aphasia in the chronic phase of LH cortical stroke recovery (average 52 months post-stroke). Before treatment, composite Fazekas scores were associated with poorer nonverbal executive function, but not with overall aphasia severity (WAB) or naming ability (Boston Naming Test) (Kaplan et al., 2001). After semantic feature analysis treatment, though, Varkanitsa et al. (2020) reported that higher pre-treatment DWMH negatively predicted the degree of language improvement. Under a similar study design, Wilmskoetter et al. (2019) did not find a direct effect of PVH on WAB AQ (n=48), but reported an indirect effect of PVH on AQ mediated by the number of intact long-range and short-range white matter fibers. Varkanitsa et al. (2020) speculate that the discrepancy between the findings of their study and those of Wilmskoetter et al. is due to differences in leukoaraiosis severity among their participants, as well as sample size differences.

Basilakos et al. (2019) highlighted what seems to be an important difference between acute and chronic aphasia as they relate to leukoaraiosis. In their sample of left MCA cortical strokes (n=35), lesion volume and time post-stroke but *not* Fazekas scores were predictors of WAB AQ during the acute phase. However, Fazekas scores at onset and initial aphasia severity together were predictors of AQ at follow-up ≥ 6 months post-stroke (mean test-retest interval was 34.8 months). The authors reported that more severe leukoaraiosis was associated with a 4.3-fold increase in the likelihood of language decline from the acute to chronic stages. If parallels in small vessel disease do exist between cortical and subcortical stroke, leukoaraiosis after subcortical stroke may be a better predictor of language outcomes in the chronic phase or in response to treatment than during the acute phase. This relationship is worth investigating in a sample of chronic subcortical stroke patients, especially those who receive treatment. The most salient difference, of course, between the present study and those cited above is our use of a strictly subcortical sample, whereas other studies deal either exclusively or primarily with cortical stroke patients. Damage to certain cortical structures is well-linked to language impairments, often reproducing classical aphasias (Hope et al., 2013). It is possible that the presence of leukoaraiosis only contributes to language deficits when there is cortical damage, but not if core language cortex is preserved (as is the case in subcortical stroke). So, while prior studies point to a link between white matter hyperintensities and language outcomes after LH stroke in general, it does not appear that this link extends to the narrower population of acute subcortical stroke patients. If leukoaraiosis severity *is* predictive of subcortical aphasia severity, our findings appear to rule out such a relationship in the acute phase.

### 4.4. Hypoperfusion

Hypoperfusion in subcortical stroke has been found to be the driving factor in acute aphasia, but not in the present study. In our sample, patients were no more likely to exhibit cortical hypoperfusion than non-aphasic patients. All three patients with poorest language performance were given an FHV score of 1/12, indicating that the presence of hypoperfusion might have contributed to their aphasia. Most participants showed little to no hypoperfusion. In our sample, 60.2% of participants received FHV ratings of 0, while 23.1% received a score of 1 (n=78). Only four participants received an FHV score of 5 or greater, and the maximum score of any participant was 7 out of a possible 12 points. Moreover, 13 of 21 aphasic patients had an FHV score of 0, and six aphasic patients received an FHV score of 1 or 2. The remaining two patients scored no higher than 6. However, it is not known if FHV rating of 1 or 2 is a sensitive measure of cortical hypoperfusion.

That the majority of our sample exhibited no cortical hypoperfusion suggests cortical hypoperfusion is not common after LH acute subcortical stroke, at least as measured by FHV. There is significant support, however, for the hypothesis that cortical hypoperfusion (measured with MRI PWI or PET) is a driver of subcortical aphasia. Olsen et al. (1986) found that in patients whose acute subcortical aphasia resolved within three months of onset (n=8), their lesions were still permanent on CT imaging. Therefore, the authors speculated that subcortical aphasia was caused by hypoperfusion instead of infarcted subcortical tissue. The resolution of language deficits, they believed, was due to enhanced blood flow to hypoperfused cortical areas, which were preserved in the meantime by collateral circulation. This hypothesis was later supported using PET imaging in acute cortical stroke (Jørgensen et al., 1994). Hillis et al. (2002) found that all subjects with aphasia (n=40) exhibited concurrent cortical hypoperfusion using time to peak (TTP) maps from MRI PWI. All six patients in this study who received interventions to restore perfusion (i.e., carotid endarterectomy, carotid stenting, or administration of pressors) immediately showed language improvements. The one aphasic patient who did not receive these interventions showed neither a relief of language deficits nor lessened cortical hypoperfusion. In a later study by Hillis et al. (2004), all 13 patients with acute subcortical aphasia exhibited cortical hypoperfusion, whereas only one non-aphasic patient (n=24) had cortical hypoperfusion (also measured from TTP maps with a similar threshold for hypoperfusion). In both studies, hypoperfusion was measured from TTP maps on PWI, where hypoperfusion is characterized by a > 2.5 second delay compared with the unaffected hemisphere. In accordance with these findings, Choi et al. (2007) found cortical hypoperfusion in all 15 of their aphasic participants. In their study, the severity of aphasia correlated positively with the extent of cortical hypoperfusion. Sebastian et al. (2014), however, found that aphasia after thalamic stroke was not due to cortical hypoperfusion, since four of the five aphasic patients in this study had cortical perfusion that was within normal limits.

The Sebastian et al. findings suggest that the role of hypoperfusion may vary depending on the location of the subcortical infarct. In the study by Hillis et al. (2004), all participants suffered basal ganglia strokes — at least to the left caudate, but in some cases also the putamen, globus pallidus, and internal capsule. Similarly, all participants in the study by Choi et al. (2007) had striatocapsular lesions. Studies whose participants suffered basal ganglia infarcts seem to implicate hypoperfusion as a driver of aphasia, whereas a sample of thalamic stroke patients implicates the infarcted region itself. This distinction may have been lost in our sample, which was quite heterogeneous; our study contained patients whose lesions were restricted to the thalamus, basal ganglia, and/or adjacent white matter structures, but no region was overrepresented as in the aforementioned studies.

Due to the limited availability of perfusion-weighted imaging in our sample, we captured cortical hypoperfusion differently than did previous studies. The qualitative nature of the FHV ratings and their lack of voxel-level specificity posed a challenge to this analysis. We did not confirm, using this approach, that the hypoperfusion is the primary driver of subcortical aphasia. Another difference between studies is that Olsen et al. (1986) and Hillis et al. (2002, 2004) used more sensitive imaging measures of cortical hypoperfusion (PET or MRI PWI). Use of FVH has not been determined to be as sensitive in detecting cortical hypoperfusion, although (when present) the number of cortical FHV correlates with the volume of hypoperfusion.

### 4.5. Patients with severe language deficits

We identified three patients who had the most severe language impairments (identifying as 1.5 SD below the sample summary score mean) in our study. A lingering question from this discussion—and one that is relevant when considering these three individuals—is whether language deficits after subcortical stroke are due to a domain-general cognitive impairment or to direct insult to linguistic processing. When language is disrupted, we might suspect domain-general functions including (but not limited to) executive function, motor function, and working memory as being affected.

All three patients discussed here suffered lesions to the basal ganglia, and one extended to the internal capsule — although none had significant thalamic involvement. Perhaps, then, we cannot shed light on the role of the thalamus, if any, in processing language directly. It is worth noting, however, that the literature appears to favor a supportive, rather than direct role for *both* the thalamus and basal ganglia in language (see Section 1). If the basal ganglia provide only computational support to language, what might we expect from ischemic lesions to its structures? A reasonable answer, write Bohsali & Crosson (2016), would be minor impairments in the accuracy or speed of word-finding rather than a severe deficit as is seen in cortical aphasia. This explanation accounts for the type and lower severity of language symptoms seen in our sample, and may also be consistent with the profiles of our three subjects with especially poor performance on language assessments. Perhaps even when subcortical strokes *do* result in aphasia, those impairments are driven by non-linguistic deficits, namely in executive functioning, working memory, and the motoric aspects of speech production.

Poor performance in Patient 1’s forward and backwards word span, as well as backward digit span, suggest a burden on working memory. Perhaps, then, Patient 1’s impaired sentence comprehension, especially for complex syntactic structures and multistep commands, is linked to his working memory deficits rather than language, per se. Next, Patient 2’s off-target word choice errors and difficulty with multistep commands may indicate deficits in domain-general executive function skills, or potentially impaired semantic processing. Although, Patient 2 had a fall at the time of his stroke, so we cannot rule out the possibility that his aphasia was due to, or exacerbated by, his fall with potential head injury. Patient 3, on the other hand, had intact comprehension but presented with severe expressive deficits. Patient 3’s poor articulatory agility and communication primarily through gestures suggest a motor speech deficit. From these cases and with support from the literature, poor language performance after acute LH subcortical stroke may not actually reflect language impairments per se. We are therefore unable to corroborate accounts explaining a non-motoric or non-executive linguistic processing role for the subcortical structures examined in this study, at least not in cases of severe language deficits.

Recall that age was the only variable that independently predicted aphasia in our study. Higher age is associated with a decline in word-retrieval and executive function, and so it seems plausible that the additional insult of a subcortical infarct may exacerbate normal age-related language decline.

### 4.6. Limitations

Several elements of this study posed a challenge to our analysis and limited comparison with previous studies. For one, our participants were assessed over the course of 17 years, and different batteries were used over the years to assess their language. Each of these batteries likely had different sensitivities to aphasia and was geared toward measuring different language functions. This necessitated the use of a summary z-score to allow comparisons across different batteries. Previous studies on subcortical aphasia often used only one battery, and were able to collect repetition data, which was not present in much of our sample. The use of different batteries also posed a challenge when comparing our results to those of previous studies. In addition, the literature does not contain an established aphasia cutoff for the BDAE, unlike the WAB-R and our in-house lexical battery (based on prior publications) (Hillis et al., 2002; Kertesz et al., 2007). Notably, we re-analyzed our data excluding participants with the BDAE, and our findings did not change.

As referenced previously, perfusion-weighted imaging and time-to-peak maps were only available for a small fraction of our study participants. Instead, we used FLAIR, which was readily available for nearly every participant, but generated a more qualitative metric for cortical hypoperfusion in the form of FHV scores. Whether our results are replicable in subjects with PWI data on record (and ideally, who received a single language battery) is an interesting avenue for further research. Similarly, the subjectivity of visual, qualitative ratings to index leukoaraiosis severity presented a challenge to this analysis, and has for other authors as well. Caligiuri et al. (2015) emphasized relatively low inter-rater reliability for qualitative visual rating scales of white matter hyperintensities. Regional differences in white matter disease burden beyond periventricular versus deep white matter hyperintensities could not be captured using the Fazekas scale, so perhaps further research using a voxel-level tool to quantify leukoaraiosis in specific structures is warranted.

## 5. Conclusion

In this study, we investigated relationships between brain structure variables and presence of aphasia in survivors of acute left hemisphere subcortical stroke. Compared to patients without aphasia, patients with aphasia had greater acute and total lesion volume, were older, and had significantly greater damage to the internal capsule (which did not survive controlling for total lesion volume). Patients with aphasia did not differ from non-aphasic patients by other demographic or stroke variables. Within a logistic regression model, only age significantly predicted aphasia status. Given the variability in language deficits and imaging markers associated with such deficits, it seems likely that subcortical aphasia is a heterogeneous clinical syndrome with distinct causes across individuals. The focus of the present study on the acute phase of LH subcortical stroke motivates further investigation into longitudinal outcomes for patients with this clinical presentation (i.e., subacute, chronic phases). Whether those results accord with the results described here may indicate which phase of stroke recovery is the most important for the characterization of deficits and interventions for addressing them.

## Data Availability

The data set generated and analyzed during the present study is publicly available in the Open Science Framework repository (https://doi.org/10.17605/OSF.IO/QYP3C).

## CRediT Authorship Contribution Statement

**Massoud Sharif:** Conceptualization, Methodology, Data Curation, Investigation, Writing – original draft, Writing – review & editing. **Emily Goldberg:** Resources, Investigation, Writing – review & editing. **Alexandra Walker:** Investigation, Writing – review & editing. **Argye Hillis:** Conceptualization, Funding acquisition, Supervision, Writing – review & editing. **Erin Meier:** Conceptualization, Methodology, Formal analysis, Writing – original draft, Writing – review & editing.

## Acknowledgements

We sincerely thank current and former members of the Stroke Cognitive Outcomes and Recovery (SCORE) Laboratory at Johns Hopkins for their support throughout the duration of this project. We also gratefully acknowledge the individuals spanning nearly two decades whose participation made this research possible.

## Abbreviations

AQ: aphasia quotient
BDAE: Boston Diagnostic Aphasia Examination
DWMH: deep white matter hyperintensity
DWI: diffusion-weighted imaging
FLAIR: fluid-attenuated inversion recovery
LB: lexical battery
MRI: magnetic resonance imaging
PVH: periventricular hyperintensity
ROI: region of interest
TTP: time-to-peak
WAB-R: Western Aphasia Battery, Revised

## References

Allan, C. M., Turner, J. W., & Gadea-Ciria, M. (1966). Investigations Into Speech Disturbances Following Stereotaxic Surgery For Parkinsonism. International Journal of Language and Communication Disorders, 1(1), 55–59.

Arba, F., Palumbo, V., Boulanger, J.-M., Pracucci, G., Inzitari, D., Buchan, A. M., Hill, M. D., & CASES Investigators. (2016). Leukoaraiosis and lacunes are associated with poor clinical outcomes in ischemic stroke patients treated with intravenous thrombolysis. International Journal of Stroke: Official Journal of the International Stroke Society, 11(1), 62–67. https://doi.org/10.1177/1747493015607517

Arsava, E. M., Rahman, R., Rosand, J., Lu, J., Smith, E. E., Rost, N. S., Singhal, A. B., Lev, M. H., Furie, K. L., Koroshetz, W. J., Sorensen, A. G., & Ay, H. (2009). Severity of leukoaraiosis correlates with clinical outcome after ischemic stroke. Neurology, 72(16), 1403–1410. https://doi.org/10.1212/WNL.0b013e3181a18823

Ay Hakan, Arsava E. Murat, Rosand Jonathan, Furie Karen L., Singhal Aneesh B., Schaefer Pamela W., Wu Ona, Gonzalez R. Gilberto, Koroshetz Walter J., & Sorensen A. Gregory. (2008). Severity of Leukoaraiosis and Susceptibility to Infarct Growth in Acute Stroke. Stroke, 39(5), 1409–1413. https://doi.org/10.1161/STROKEAHA.107.501932

Bahrainwala, Z. S., Hillis, A. E., Dearborn, J., & Gottesman, R. F. (2014). Neglect performance in acute stroke is related to severity of white matter hyperintensities. Cerebrovascular Diseases (Basel, Switzerland), 37(3), 223–230. https://doi.org/10.1159/000357661

Basilakos, A., Stark, B. C., Johnson, L., Rorden, C., Yourganov, G., Bonilha, L., & Fridriksson, J. (2019). Leukoaraiosis Is Associated With a Decline in Language Abilities in Chronic Aphasia. Neurorehabilitation and Neural Repair, 33(9), 718–729. https://doi.org/10.1177/1545968319862561

Basile, A. M., Pantoni, L., Pracucci, G., Asplund, K., Chabriat, H., Erkinjuntti, T., Fazekas, F., Ferro, J. M., Hennerici, M., O’Brien, J., Scheltens, P., Visser, M. C., Wahlund, L.-O., Waldemar, G., Wallin, A., Inzitari, D., & LADIS Study Group. (2006). Age, hypertension, and lacunar stroke are the major determinants of the severity of age-related white matter changes. The LADIS (Leukoaraiosis and Disability in the Elderly) Study. Cerebrovascular Diseases (Basel, Switzerland), 21(5–6), 315–322. https://doi.org/10.1159/000091536

Berthier, M. L. (2005). Poststroke aphasia: Epidemiology, pathophysiology and treatment. Drugs & Aging, 22(2), 163–182. https://doi.org/10.2165/00002512-200522020-00006

Bogousslavsky, J., & Caplan, L. R. (1993). Vertebrobasilar Occlusive Disease: Review of Selected Aspects. Cerebrovascular Diseases, 3(4), 193–205. https://doi.org/10.1159/000108701

Bohsali, A., & Crosson, B. (2016). The basal ganglia and language: A tale of two loops. In The basal ganglia: Novel perspectives on motor and cognitive functions (pp. 217–242).Springer International Publishing. https://doi.org/10.1007/978-3-319-42743-0_10

Bouvier, L., Groulx, B., Martel-Sauvageau, V., & Monetta, L. (2017). Language disturbances after non-thalamic subcortical stroke: A review of the literature. Geriatrie Et Psychologie Neuropsychiatrie Du Vieillissement, 15(2), 173–184. https://doi.org/10.1684/pnv.2017.0666

Broadbent, W. H. (1872). On the Cerebral Mechanism of Speech and Thought. Medico-Chirurgical Transactions, 55, 145–194.

Caligiuri, M. E., Perrotta, P., Augimeri, A., Rocca, F., Quattrone, A., & Cherubini, A. (2015). Automatic Detection of White Matter Hyperintensities in Healthy Aging and Pathology Using Magnetic Resonance Imaging: A Review. Neuroinformatics, 13(3), 261–276. https://doi.org/10.1007/s12021-015-9260-y

Caplan, L. R., Schmahmann, J. D., Kase, C. S., Feldmann, E., Baquis, G., Greenberg, J. P., Gorelick, P. B., Helgason, C., & Hier, D. B. (1990). Caudate Infarcts. Archives of Neurology, 47(2), 133–143. https://doi.org/10.1001/archneur.1990.00530020029011

Choi, J. Y., Lee, K. H., Na, D. L., Byun, H. S., Lee, S. J., Kim, H., Kwon, M., Lee, K.-H., & Kim, B.-T. (2007). Subcortical aphasia after striatocapsular infarction: Quantitative analysis of brain perfusion SPECT using statistical parametric mapping and a statistical probabilistic anatomic map. Journal of Nuclear Medicine: Official Publication, Society of Nuclear Medicine, 48(2), 194–200.

Crosson, B. (1984). Role of the dominant thalamus in language: A review. Psychological Bulletin, 96(3), 491–517. https://doi.org/10.1037/0033-2909.96.3.491

Crosson, B. (1985). Subcortical functions in language: A working model. Brain and Language, 25(2), 257–292. https://doi.org/10.1016/0093-934x(85)90085-9

Crosson, B. (2019). The Role of Cortico-Thalamo-Cortical Circuits in Language: Recurrent Circuits Revisited. Neuropsychology Review, 31(3), 516–533. https://doi.org/10.1007/s11065-019-09421-8

Crosson, B., Moore, A. B., Gopinath, K., White, K. D., Wierenga, C. E., Gaiefsky, M. E., Fabrizio, K. S., Peck, K. K., Soltysik, D., Milsted, C., Briggs, R. W., Conway, T. W., & Rothi, L. J. G. (2005). Role of the Right and Left Hemispheres in Recovery of Function during Treatment of Intention in Aphasia. Journal of Cognitive Neuroscience, 17(3), 392–406. https://doi.org/10.1162/0898929053279487

Damasio, A. R., Damasio, H., Rizzo, M., Varney, N., & Gersh, F. (1982). Aphasia with nonhemorrhagic lesions in the basal ganglia and internal capsule. Archives of Neurology, 39(1), 15–24. https://doi.org/10.1001/archneur.1982.00510130017003

De Witte, L., Brouns, R., Kavadias, D., Engelborghs, S., De Deyn, P. P., & Mariën, P. (2011). Cognitive, affective and behavioural disturbances following vascular thalamic lesions: A review. Cortex; a Journal Devoted to the Study of the Nervous System and Behavior, 47(3), 273–319. https://doi.org/10.1016/j.cortex.2010.09.002

Fazekas, F., Chawluk, J. B., Alavi, A., Hurtig, H. I., & Zimmerman, R. A. (1987). MR signal abnormalities at 1.5 T in Alzheimer’s dementia and normal aging. AJR. American Journal of Roentgenology, 149(2), 351–356. https://doi.org/10.2214/ajr.149.2.351

Fierini, F., Poggesi, A., & Pantoni, L. (2017). Leukoaraiosis as an outcome predictor in the acute and subacute phases of stroke. Expert Review of Neurotherapeutics, 17(10), 963–975. https://doi.org/10.1080/14737175.2017.1371013

Flowers, H. L., Skoretz, S. A., Silver, F. L., Rochon, E., Fang, J., Flamand-Roze, C., & Martino, R. (2016). Poststroke Aphasia Frequency, Recovery, and Outcomes: A Systematic Review and Meta-Analysis. Archives of Physical Medicine and Rehabilitation, 97(12), 2188-2201.e8. https://doi.org/10.1016/j.apmr.2016.03.006

Goldberg, E. B., Meier, E. L., Sheppard, S. M., Breining, B. L., & Hillis, A. E. (2021). Stroke Recurrence and Its Relationship With Language Abilities. Journal of Speech, Language, and Hearing Research: JSLHR, 64(6), 2022–2037. https://doi.org/10.1044/2021_JSLHR-20-00347

Goodglass, H., Kaplan, E., & Barresi, B. (2001a). BDAE-3: Boston Diagnostic Aphasia Examination—Third Edition. Lippincott Williams & Wilkins.

Goodglass, H., Kaplan, E., & Barresi, B. (2001b). The Assessment of Aphasia and Related Disorders. Lippincott Williams & Wilkins.

Henninger, N., Khan, M. A., Zhang, J., Moonis, M., & Goddeau, R. P. (2014). Leukoaraiosis predicts cortical infarct volume after distal middle cerebral artery occlusion. Stroke, 45(3), 689–695. https://doi.org/10.1161/STROKEAHA.113.002855

Henninger, N., Lin, E., Haussen, D. C., Lehman, L. L., Takhtani, D., Selim, M., & Moonis, M. (2013). Leukoaraiosis and sex predict the hyperacute ischemic core volume. Stroke, 44(1), 61–67. https://doi.org/10.1161/STROKEAHA.112.679084

Hillis, A. E., Barker, P. B., Wityk, R. J., Aldrich, E. M., Restrepo, L., Breese, E. L., & Work, M. (2004). Variability in subcortical aphasia is due to variable sites of cortical hypoperfusion. Brain and Language, 89(3), 524–530. https://doi.org/10.1016/j.bandl.2004.01.007

Hillis, A. E., Kleinman, J. T., Newhart, M., Heidler-Gary, J., Gottesman, R., Barker, P. B., Aldrich, E., Llinas, R., Wityk, R., & Chaudhry, P. (2006). Restoring Cerebral Blood Flow Reveals Neural Regions Critical for Naming. The Journal of Neuroscience, 26(31), 8069–8073. https://doi.org/10.1523/JNEUROSCI.2088-06.2006

Hillis, A. E., Wityk, R. J., Barker, P. B., Beauchamp, N. J., Gailloud, P., Murphy, K., Cooper, O., & Metter, E. J. (2002). Subcortical aphasia and neglect in acute stroke: The role of cortical hypoperfusion. Brain, 125(5), 1094–1104. https://doi.org/10.1093/brain/awf113

Hope, T. M. H., Seghier, M. L., Leff, A. P., & Price, C. J. (2013). Predicting outcome and recovery after stroke with lesions extracted from MRI images. NeuroImage. Clinical, 2, 424–433. https://doi.org/10.1016/j.nicl.2013.03.005

Jørgensen, H. S., Sperling, B., Nakayama, H., Raaschou, H. O., & Olsen, T. S. (1994). Spontaneous reperfusion of cerebral infarcts in patients with acute stroke. Incidence, time course, and clinical outcome in the Copenhagen Stroke Study. Archives of Neurology, 51(9), 865–873. https://doi.org/10.1001/archneur.1994.00540210037011

Kang, E. K., Sohn, H. M., Han, M.-K., & Paik, N.-J. (2017). Subcortical Aphasia After Stroke. Annals of Rehabilitation Medicine, 41(5), 725–733. https://doi.org/10.5535/arm.2017.41.5.725

Kaplan, E., Goodglass, H., Weintraub, S., Segal, O., & Loon-Vervoorn A. van. (2001). Boston naming test. Pro-ed.

Karussis, D., Leker, R. R., & Abramsky, O. (2000). Cognitive dysfunction following thalamic stroke: A study of 16 cases and review of the literature. Journal of the Neurological Sciences, 172(1), 25–29. https://doi.org/10.1016/s0022-510x(99)00267-1

Kennedy, M., & Murdoch, B. E. (1993). Chronic Aphasia Subsequent to Striato-capsular and Thalamic Lesions in the Left Hemisphere. Brain and Language, 44(3), 284–295. https://doi.org/10.1006/brln.1993.1019

Kertesz, A., Kertesz, A., Raven, J. C., & PsychCorp (Firm). (2007). WAB-R: Western Aphasia Battery-Revised. PsychCorp.

Kuljic-Obradovic, D. C. (2003). Subcortical aphasia: Three different language disorder syndromes? European Journal of Neurology, 10(4), 445–448. https://doi.org/10.1046/j.1468-1331.2003.00604.x

Kussmaul, A. (1877). Disturbances of speech. In Cyclopedie of the practice of medicine (Vol. 14). William Wood.

Llano, D. A. (2013). Functional imaging of the thalamus in language. Brain and Language, 126(1), 62–72. https://doi.org/10.1016/j.bandl.2012.06.004

Manolio, T. A., Kronmal, R. A., Burke, G. L., Poirier, V., O’Leary, D. H., Gardin, J. M., Fried, L. P., Steinberg, E. P., & Bryan, R. N. (1994). Magnetic resonance abnormalities and cardiovascular disease in older adults. The Cardiovascular Health Study. Stroke, 25(2), 318–327. https://doi.org/10.1161/01.str.25.2.318

Marek, M., Horyniecki, M., Frączek, M., & Kluczewska, E. (2018). Leukoaraiosis – new concepts and modern imaging. Polish Journal of Radiology, 83, e76–e81. https://doi.org/10.5114/pjr.2018.74344

Mega, M. S., & Alexander, M. P. (1994). Subcortical aphasia: The core profile of capsulostriatal infarction. Neurology, 44(10), 1824–1829. https://doi.org/10.1212/wnl.44.10.1824

Murdoch, B. E. (2001). Subcortical brain mechanisms in speech and language. Folia Phoniatrica et Logopaedica: Official Organ of the International Association of Logopedics and Phoniatrics (IALP), 53(5), 233–251. https://doi.org/10.1159/000052679

Nadeau, S. E. (2021). Basal Ganglia and Thalamic Contributions to Language Function: Insights from A Parallel Distributed Processing Perspective. Neuropsychology Review, 31(3), 495– 515. https://doi.org/10.1007/s11065-020-09466-0

Nadeau, S. E., & Crosson, B. (1997). Subcortical Aphasia. Brain and Language, 58(3), 355–402. https://doi.org/10.1006/brln.1997.1707

NITRC: NiiStat. (n.d.). NeuroImaging Tools & Resources Collaboratory. Retrieved January 6, 2022, from https://www.nitrc.org/projects/niistat/

Olsen, T. S., Bruhn, P., & Oberg, R. G. (1986). Cortical hypoperfusion as a possible cause of “subcortical aphasia.” Brain: A Journal of Neurology, 109 (Pt 3), 393–410. https://doi.org/10.1093/brain/109.3.393

Paradise, M. B., Shepherd, C. E., Wen, W., & Sachdev, P. S. (2018). Neuroimaging and neuropathology indices of cerebrovascular disease burden: A systematic review. Neurology, 91(7), 310–320. https://doi.org/10.1212/WNL.0000000000005997

Payabvash, S., Souza, L. C., Wang, Y., Schaefer, P. W., Furie, K. L., Halpern, E. F., Gonzalez, R. G., & Lev, M. H. (2011). Regional Ischemic Vulnerability of the Brain to Hypoperfusion: The Need for Location Specific CT Perfusion Thresholds in Acute Stroke Patients. Stroke; a Journal of Cerebral Circulation, 42(5), 1255–1260. https://doi.org/10.1161/STROKEAHA.110.600940

Plowman, E., Hentz, B., & Ellis, C. (2012). Post-stroke aphasia prognosis: A review of patient-related and stroke-related factors. Journal of Evaluation in Clinical Practice, 18(3), 689– 694. https://doi.org/10.1111/j.1365-2753.2011.01650.x

Radanovic, M., & Mansur, L. L. (2017). Aphasia in vascular lesions of the basal ganglia: A comprehensive review. Brain and Language, 173, 20–32. https://doi.org/10.1016/j.bandl.2017.05.003

Rangus, I., Fritsch, M., Endres, M., Udke, B., & Nolte, C. H. (2021). Frequency and phenotype of thalamic aphasia. Journal of Neurology. https://doi.org/10.1007/s00415-021-10640-4

Reyes Dennys, Hitomi Emi, Simpkins Alexis, Lynch John, Hsia Amie, Benson Richard, Nadareishvili Zurab, Luby Marie, Latour Lawrence, & Leigh Richard. (2017). Abstract TP63: Detection of Perfusion Deficits Using FLAIR and GRE Based Vessel Signs. Stroke, 48(suppl_1), ATP63–ATP63. https://doi.org/10.1161/str.48.suppl_1.tp63

Rorden, C., & Brett, M. (2000). Stereotaxic display of brain lesions. Behavioural Neurology, 12(4), 191–200. https://doi.org/10.1155/2000/421719

Sambin, S., Teichmann, M., de Diego Balaguer, R., Giavazzi, M., Sportiche, D., Schlenker, P., & Bachoud-Lévi, A.-C. (2012). The role of the striatum in sentence processing: Disentangling syntax from working memory in Huntington’s disease. Neuropsychologia, 50(11), 2625–2635. https://doi.org/10.1016/j.neuropsychologia.2012.07.014

Sebastian, R., Schein, M. G., Davis, C., Gomez, Y., Newhart, M., Oishi, K., & Hillis, A. E. (2014). Aphasia or Neglect after Thalamic Stroke: The Various Ways They may be Related to Cortical Hypoperfusion. Frontiers in Neurology, 5, 231. https://doi.org/10.3389/fneur.2014.00231

Shi, E. R., & Zhang, Q. (2020). A domain-general perspective on the role of the basal ganglia in language and music: Benefits of music therapy for the treatment of aphasia. Brain and Language, 206, 104811. https://doi.org/10.1016/j.bandl.2020.104811

Svennilson, E., Torvik, A., Lowe, R., & Leksell, L. (1960). TREATMENT OF PARKINSONISM BY STEREOTACTIC THERMOLESIONS IN THE PALLIDAL REGION. A clinical evaluation of 81 cases. Acta Psychiatrica Scandinavica, 35(3), 358–377. https://doi.org/10.1111/j.1600-0447.1960.tb07606.x

Te, M., Zhao, E., Zheng, X., Sun, Q., & Qu, C. (2015). Leukoaraiosis with mild cognitive impairment. Neurological Research, 37(5), 410–414. https://doi.org/10.1179/1743132815Y.0000000028

Tsouli, S., Kyritsis, A. P., Tsagalis, G., Virvidaki, E., & Vemmos, K. N. (2009). Significance of aphasia after first-ever acute stroke: Impact on early and late outcomes. Neuroepidemiology, 33(2), 96–102. https://doi.org/10.1159/000222091

Varkanitsa, M., Peñaloza, C., Charidimou, A., Caplan, D., & Kiran, S. (2020). White Matter Hyperintensities Predict Response to Language Treatment in Poststroke Aphasia. Neurorehabilitation and Neural Repair, 34(10), 945–953. https://doi.org/10.1177/1545968320952809

Vedala, K., Nagabandi, A. K., Looney, S., & Bruno, A. (2019). Factors Associated with Leukoaraiosis Severity in Acute Stroke Patients. Journal of Stroke and Cerebrovascular Diseases: The Official Journal of National Stroke Association, 28(7), 1897–1901. https://doi.org/10.1016/j.jstrokecerebrovasdis.2019.04.003

Wahlund, L. O., Barkhof, F., Fazekas, F., Bronge, L., Augustin, M., Sjögren, M., Wallin, A., Ader, H., Leys, D., Pantoni, L., Pasquier, F., Erkinjuntti, T., Scheltens, P., & European Task Force on Age-Related White Matter Changes. (2001). A new rating scale for age-related white matter changes applicable to MRI and CT. Stroke, 32(6), 1318–1322. https://doi.org/10.1161/01.str.32.6.1318

Wernicke, C. (1874). Der aphasische Symptomenkomplex. In C. Wernicke (Ed.), Der aphasische Symptomencomplex: Eine psychologische Studie auf anatomischer Basis (pp. 1–70). Springer. https://doi.org/10.1007/978-3-642-65950-8_1

Wilmskoetter, J., Marebwa, B., Basilakos, A., Fridriksson, J., Rorden, C., Stark, B. C., Johnson, L., Hickok, G., Hillis, A. E., & Bonilha, L. (2019). Long-range fibre damage in small vessel brain disease affects aphasia severity. Brain, 142(10), 3190–3201. https://doi.org/10.1093/brain/awz251

Wright, A., Tippett, D., Saxena, S., Sebastian, R., Breining, B., Faria, A., & Hillis, A. E. (2018). Leukoaraiosis is independently associated with naming outcome in poststroke aphasia. Neurology, 91(6), e526–e532. https://doi.org/10.1212/WNL.0000000000005945

Yassi, N., Churilov, L., Campbell, B. C. V., Sharma, G., Bammer, R., Desmond, P. M., Parsons, M. W., Albers, G. W., Donnan, G. A., Davis, S. M., EPITHET Investigators, & DEFUSE Investigators. (2015). The association between lesion location and functional outcome after ischemic stroke. International Journal of Stroke: Official Journal of the International Stroke Society, 10(8), 1270–1276. https://doi.org/10.1111/ijs.12537

Yuan, J., Feng, L., Hu, W., & Zhang, Y. (2018). Use of Multimodal Magnetic Resonance Imaging Techniques to Explore Cognitive Impairment in Leukoaraiosis. Medical Science Monitor : International Medical Journal of Experimental and Clinical Research, 24, 8910–8915. https://doi.org/10.12659/MSM.912153

