## Supplemental Tables for "The contribution of white matter pathology, hypoperfusion, lesion load, and stroke recurrence to language deficits following acute subcortical left hemisphere stroke"

| Western Aphasia Battery - Revised (WAB-R) | Boston Diagnostic Aphasia Examination (BDAE) | Lexical Battery (LB) |
|---|---|---|
| Information content | Simple social responses | Oral naming |
| Fluency, grammatical competence | Word comprehension | Tactile naming |
| Yes/no questions | Complex ideational material | Auditory comprehension |
| Auditory word recognition | Automatized sequences | |
| Sequential commands | Responsive naming | |
| Object naming | | |
| Word fluency | | |
| Sentence completion | | |
| Responsive speech | | |

**Supplemental Table 1. Subtests taken from each language battery.** Subtests from the Western Aphasia Battery – Revised (WAB-R), Boston Diagnostic Aphasia Examination (BDAE), and an in-house lexical battery (LB) were chosen to reflect auditory comprehension, naming, and verbal expression skills.

**A. Patient 1**

| Western Aphasia Battery | |
|---|---|
| Information content | 0/10 |
| Fluency, grammatical competence | 0/10 |
| Yes/no questions | 60/60 |
| Auditory word recognition | 54/60 |
| Sequential commands | 40/80 |
| Repetition | 0/100 |
| Object naming | 0/60 |
| Word fluency | 0/20 |
| Sentence completion | 0/10 |
| Responsive speech | 0/10 |
| *Aphasia quotient* | *55.4/100* |

| Additional Tests | |
|---|---|
| Digit span, forward | 5 |
| Digit span, backward | 0 |
| Word span, forward | 3 |
| Word span, backward | 0 |
| Sentence-picture matching (% correct) | 71.25 |
| Enactment of spoken sentences (% correct) | 73.75 |

**B. Patient 2**

| Western Aphasia Battery | |
|---|---|
| Information content | 8/10 |
| Fluency, grammatical competence | 7/10 |
| Yes/no questions | 51/60 |
| Auditory word recognition | 53/60 |
| Sequential commands | 55/80 |
| Repetition | 100/100 |
| Object naming | 42/60 |
| Word fluency | 1/20 |
| Sentence completion | 8/10 |
| Responsive speech | 8/10 |
| *Aphasia quotient* | *76.9/100* |

| Apraxia Battery for Adults | |
|---|---|
| Increasing word length, Part A (deterioration in performance) | 0/20 |
| Increasing word length, Part B (deterioration in performance) | 1/20 |
| Repeated trials | 29/30 |
| Inventory of articulation (errors) | 2/15 |

| Additional Tests | |
|---|---|
| Digit span, forward | 6 |
| Digit span, backward | 5 |
| Word span, forward | 4 |
| Word span, backward | 0 |
| Thematic role assignment, video sentences | 47/80 |
| Thematic role assignment, synonym judgment (semantic condition) | 23/40 |
| Thematic role assignment, synonym judgment (syntactic condition) | 23/40 |

**C. Patient 3**

| Boston Diagnostic Aphasia Examination | |
|---|---|
| Simple social responses | 2/7 |
| Severity of speech output profile (0-5) | 1 |
| Word comprehension | 15/16 |
| Complex ideational material | 5/6 |
| Automatized sequences | 0/4 |
| Single word repetition | 2/5 |
| Sentence repetition | 0/2 |
| Responsive naming | 0/10 |
| Screening of special categories | 5/12 |
| Basic symbol recognition | 4/4 |
| Number matching | 4/4 |
| Word identification, picture-word matching | 4/4 |
| Basic oral word reading (points) | 0 |
| Oral reading of sentences (# correct) | 0 |
| Well-formedness of written letters | 4/14 |
| Correctness of written letter choice | 12/21 |
| Motor facility | 4/14 |

| Rating Scale Profile of Speech Characteristics | |
|---|---|
| Articulatory agility | 2/7 |
| Phrase length | 1/7 |
| Grammatical form | 1/7 |
| Sentence repetition (percentile range) | 0-20 |
| Auditory comprehension (percentile range) | 90-100 |

| Additional Tests | |
|---|---|
| Pyramids & Palm Trees | 15/15 |
| Kissing and Dancing | 7/15 |

**Supplemental Table 2. Language data from three patients with severe language deficits.** Results from additional behavioral tasks are also included where relevant. (A) Patient 1 received the Western Aphasia Battery – Revised (WAB-R) and additional tests for word span, digit span, and comprehending movement-derived sentences. This patient was non-verbal, and used a communication board. (B) Patient 2 received the WAB-R, the Apraxia Battery for Adults, and additional tests for digit span, word span, and thematic role assignment. (C) Patient 3 received the Boston Diagnostic Aphasia Examination (BDAE), which included a rating scale for profiling speech characteristics. Additional tests included Pyramids & Palm Trees as well as Kissing and Dancing.
